# Availability and Affordability of Essential Medicines from the WHO Global Core List in Albania: A Cross-Sectional Survey Using the WHO/HAI Methodology

**DOI:** 10.1101/2025.08.01.25332601

**Authors:** E. Petro, A.C.M Siebers, A.K. Mantel-Teeuwisse, F. Suleman, H.A. van den Ham

## Abstract

There is limited information on the current state of access to essential medicines in Albania. Therefore, this study investigated the accessibility of essential medicines using an adapted version of the World Health Organization/Health Action International (WHO/HAI) methodology. A cross-sectional survey of 30 private pharmacies was undertaken, across six urban areas, and for 14 medicines from the Global Core List (GCL). Data on availability, patient prices, and affordability were collected, with affordability measured using the daily wage of the lowest-paid government worker (LPGW) and the median price ratio (MPR). MPRs were calculated to compare local prices with international reference and regional benchmarks.

The mean availability of the GCL medicines was 79.8%, falling short of the WHO’s 80% benchmark, though it improved to 83.3% when availability of alternative formulations were included. Ceftriaxone, amoxicillin, and salbutamol were among the least available medicines and were frequently reported as out of stock. All medicines, except ceftriaxone, met the LPGW affordability criteria. MPR analysis showed that while most medicines were priced competitively, some, like diclofenac and ceftriaxone, were significantly more expensive than international reference prices. Price comparisons indicated that Albanian medicine prices were generally lower than those in selected Balkan and EU countries, although MPRs for five medicines were 4 times higher compared to international reference prices.

Overall, 64% of the surveyed medicines met both availability and affordability thresholds. It was found that access to medicines in Albania is primarily limited by availability rather than affordability. The study highlights significant gaps in the availability of key antibiotics and asthma medications, emphasizing the need for policy reforms to improve rational use and other pharmaceutical value chain interventions. These results are important for shaping Albania’s health policy in advancing towards equal access to essential medicines

## Introduction

Access to medicines is a multifaceted concept encompassing availability, affordability, accessibility, and acceptability. Availability pertains to the presence of required medicines in healthcare facilities, while affordability refers to the patient’s financial feasibility of obtaining these medicines. Accessibility involves the geographic proximity of healthcare outlets, and acceptability reflects the extent to which medicines meet user expectations in terms of effectiveness, quality, and suitability [1,2]. In low– and middle-income countries (LMICs), barriers to access to essential medicines remain pervasive, undermining efforts to improve public health and reduce disease burden [3].

Albania, an upper-middle-income country (UMIC) is located in Southeast Europe. With a population of approximately 2.36 million and an average per capita healthcare expenditure of $464 per year [4, 5]. Albania’s health system faces significant challenges in delivering equitable access to essential medicines. This is particularly evident in the pharmaceutical sector, where the majority of services are delivered by the private sector [6,7], through the Albanian Mandatory Health Insurance Fund (AMHIF), which was established in 1995. This agency is funded by the government and covers individuals, including minors under 18 years of age, students up to the 25 years old, pensioners and individuals receiving unemployment, social assistance or disability benefits [6]. In Albania, non-communicable diseases (NCDs) are the predominant cause of mortality, accounting for approximately 96% of all deaths in 2018, with cardiovascular diseases alone responsible for 54% of these deaths [8]. Antimicrobial resistance (AMR) is also a significant concern; in 2019, there were 339 deaths directly attributable to AMR and an additional 1,300 associated deaths [9].

The services provided by AMHIF encompass public healthcare centers, privately contracted primary care centers, tertiary hospitals and contracted pharmacies. Under this arrangement, contracted pharmacies reimburse AMHIF listed medicines, subject to a referral process through primary and secondary healthcare centers, where specialist physicians assess whether patients meet the necessary criteria and for more complex cases final decisions are made at the tertiary healthcare center, the central University hospital center in Tirana “Mother Teresa” (QSUT). If these criteria are met, medicines listed in the AMHIF reimbursement list may qualify for government reimbursement [10]. Although the reimbursement scheme is available to the entire population, it does not cover all medical conditions, and the reimbursement rates vary depending on the class of medicine [11].

The World Health Organization/Health Action International (WHO/HAI) methodology, developed in 2000, provides a standardized approach for evaluating the availability, affordability, and pricing of essential medicines in healthcare systems worldwide. It includes a Global Core List (GCL) of 14 medicines targeting prevalent diseases, and serves as a benchmark for cross-country comparisons [12]. Although the methodology has been widely applied in other LMICs, its application in middle-income countries in Europe, including Albania, has been limited representing a significant gap in the literature. As a European Union (EU) candidate country, Albania is required to align its healthcare system with EU standards, including the provision of access to essential medicines as outlined in Chapter 28 of the EU acquis [7].

Detailed health-related indicators in Table 1 show that Albania underperforms compared to EU averages across all key health and socioeconomic metrics. Life expectancy is lower for both males (70.7 vs. 77.9 years) and females (77.1 vs. 83.3 years), while neonatal mortality is more than three times higher (7 vs. 2 per 1,000 live births). Health spending is also considerably lower (6.2% vs. 10.4% of GDP). Although gaps in unemployment and poverty are narrower, the overall pattern reflects systemic weaknesses. These disparities may indicate suboptimal access to essential medicines, yet no comprehensive study has assessed the situation. Therefore, it is imperative to assess the availability and affordability of essential medicines, particularly those included in the WHO GCL, within Albania’s healthcare system using the WHO/HAI methodology. Thus, this study aims to determine if the accessibility of WHO GCL medicines in Albania will exceed 80%, in accordance with the WHO guidelines for ensuring optimal access to essential medicines.

**Table 1.** Health-related indicators in Albania compared to the EU average 115-27)

| Indicator | Albania | European Union (average) |
| --- | --- | --- |
| Gross Domestic Product per capita (in US\$) 2023 | 8,575.2 | 41,422.8 |
| Current healthcare expenditure as percentage of the Gross Domestic Product (in %) 2022 | 6.19 | 10.4 |
| Percentage of a country's population living on less than \$3.65 per day, adjusted for purchasing power parity (PPP) based on 2017 prices. 2020* | 2 | 0.6 |
| Universal Health Coverage service coverage (in %) 2021 | 64 | 83.4 |
| Unemployment, total (% of total labor force) | 8.8# | 5.7^ |
| Life expectancy males (in years) | 70.7 | 77.9 |
| Life expectancy females (in years) | 77.1 | 83.3 |
| Neonatal mortality rate (per 1000 live births) 2023 | 7 | 2 |
| Death rate, crude (per 1000 people) 2021 | 11.1 | 10.1 |

\*At Risk of Poverty or Social Exclusion (AROPE) indicator is a key metric used by the European Union (EU) to monitor social inclusion and assess the effectiveness of poverty reduction strategies. # Q4 2024.
^Feb 2025

## Materials and methods

### Study Design and Setting

A cross-sectional survey was conducted to assess the availability, prices, and affordability of medicines in Albania, using an adapted version of the WHO/HAI methodology [12]. Six survey areas were selected based on population density and proximity to Tirana, the main urban center. These included Tirana, Durrës, Fier, Vlorë, Kavajë, and Lushnjë. In each area, five private pharmacies were selected to be surveyed, with five alternates identified based on the list of pharmacies provided by the Albanian Order of Pharmacists. All surveyed facilities were privately owned and categorized as either contracted or uncontracted by the AMHIF. Contracted pharmacies are authorized to dispense reimbursed medicines to patients with valid reimbursement prescriptions, reducing out-of-pocket costs. Primary healthcare centers were excluded from the survey due to their limited list of medicines as they are only authorized to provide the emergency medicines outlined in the basic package of primary healthcare services [28].

### Medicine Selection

The survey followed the WHO/HAI methodology to ensure international comparability [12]. From the WHO GCL, 12 out of 14 medicines are included in the AMHIF list, while all the medicines are registered in Albania [11,29]. The selected medicines were validated for exact alignment with the reimbursement list of the Albanian Mandatory Health Insurance Fund (AMHIF) (see Supplementary Table S1). In cases where the standard formulations (diclofenac 50 mg tablets/capsules and co-trimoxazole 8+40 mg/mL oral suspension) were not available at surveyed facilities, alternative formulations such as diclofenac (Voltfast 50 mg powder for oral suspension) and co-trimoxazole (400/80 mg tablets) were included as potential therapeutic alternatives.

#### Ethics approval and consent to participate

This study involved human participants and Institutional approval was obtained from the Science-Geosciences Ethics Review Board (SG ERB) of Utrecht University (Sci R-23.011) and Albanian Order of Pharmacists (Protocol nr.948 date 11.12.2023). Participants were required to sign informed consent forms before the survey. Participation was voluntary and participants had the option to withdraw from the study at any point.

### Data Collection

Data collection occurred between March 7^th^ and 16^th^ 2024. Four trained data collectors, two pharmacy students, one medical student, and one pharmacist, used the KoboCollect mobile application for standardized data entry [30]. For each medicine, data on formulation, strength, price, expiry date, packaging, and availability were recorded. Both originator brands (OB) and lowest-priced generics (LPG) were included. Photos of medicine packages were taken for verification processes.

### Data Management and Analysis

Collected data were exported to Excel for cleaning and validation. Discrepancies were resolved using photo verification. Random cross-checks were conducted by the researchers to verify the accuracy of the data.

#### Determination of Availability

Availability was calculated as the percentage of pharmacies with each medicine available on the day of data collection, and was analyzed overall, by medicine type (OB/LPG), and by survey area. Availability was categorized as: not available (0%), very low (<30%), low (30–49%), fairly high (50–80%), and high (>80%) [10]. A sub-analysis was conducted to include “similar formulations”, defined as alternative dosage forms with equivalent therapeutic use (e.g., diclofenac oral suspension in place of tablets, or co-trimoxazole 400/80 mg tablets in place of pediatric-specific formulations), to provide additional insight into potential therapeutic options in the absence of age-appropriate products (see Supplementary Table S2).

#### Determination of Affordability

Affordability was assessed by considering both the day’s wages of the lowest paid governmental worker (LPGW) [12], and the median price ratio (MPR). In the WHO/HAI methodology affordability is calculated as the number of days’ wages required to purchase a full treatment course. The course of treatment refers to the total quantity of a medicine required to complete a full therapeutic regimen for a specific condition, based on the standard dosage, frequency, and duration of treatment as recommended in standard treatment guidelines (e.g. 30 days for chronic treatment). The price per treatment is found by multiplying the median unit price of the medicine by the quantity needed. The LPGW’s monthly salary was 40,000 Albanian Lek (ALL) (≈421.83 USD), equating to ∼1,430 ALL/day [31]. Affordability was determined by dividing the price per treatment by the LPGW. A medicine was deemed unaffordable if it exceeded one day’s wage, according to the WHO/HAI methodology [12].

#### Median Price Ratio

The Management Sciences for Health (MSH) international reference prices (IRPs), last published in 2015, were used to calculate the Median Price Ratio (MPR). These IRPs reflect procurement prices paid by governments and non-profit organizations in low– and middle-income countries and are recommended by WHO/HAI as reliable external benchmarks for assessing affordability [32]. To approximate current values, the 2015 IRPs were adjusted using the average Consumer Price Index (CPI) of 48 European countries (see Table S3) [33].

In addition to using IRPs, unit prices from selected comparator countries were included to contextualize local pricing. These countries comprised Croatia, Cyprus, and Slovenia, as well as early EU member states such as Belgium, France, the Netherlands, Italy, and Denmark. These prices, updated using national CPIs and converted to Albanian lek (ALL), were drawn from sources that included retail, wholesale, and unspecified price types [34,35].

MPRs were calculated by dividing the median local unit price by the corresponding IRP, in accordance with WHO/HAI methodology. Although WHO/HAI does not prescribe a fixed threshold, an MPR greater than 4.0 for generics is often considered indicative of affordability issues. Median values were used to reduce the influence of outliers and better represent typical price levels [12].

#### Stock-Outs

Stock-outs were recorded when a medicine was unavailable at the time of visit. Durations were categorized as: unavailable within the last week, last month, over a month, or never available. Medicines not stocked due to low demand were excluded. If either the originator brand (OB) or lowest-priced generic (LPG) was available, the medicine was not considered out of stock. While medicine availability was verified through on-site checking, information on stock-outs was based solely on pharmacists’ self-reports.

#### Overall Accessibility

A scatter plot illustrated the relationship between availability and affordability (LPGW method) for each medicine. Medicines available and affordable in >80% of surveyed facilities were considered accessible.

## Results

Among the initially 30 selected pharmacies, 12 pharmacies were unable to participate due to issues such as unclear schedule agreements, busy schedules of pharmacists, and refusal to participate in the study. Instead, 12 back-up pharmacies were visited within the same survey areas for a total sample of 30. An overview of the participating pharmacies and their contract status with AMHIF is presented in Table 2.

**Table 2.** Number of included pharmacies, both contracted and uncootracted.

| Cities | Durrës | Kavajë | Tirana | Vlorë | Fier | Lushnjë | Total |
| --- | --- | --- | --- | --- | --- | --- | --- |
| County | Durrës | Tirana | Tirana | Vlorë | Fier | Fier |  |
| Number of Contracted Pharmacies | 3 | 4 | 1 | 2 | 3 | 3 | 16 |
| Number of Uncontracted Pharmacies | 2 | 1 | 4 | 3 | 2 | 2 | 14 |

### Availability of Essential Medicines

The overall availability of essential medicines from the WHO GCL in surveyed pharmacies averaged 79.8%, narrowly missing the WHO’s 80% benchmark. Ceftriaxone, ciprofloxacin, metformin, and paracetamol were consistently available across all outlets. In contrast, co-trimoxazole showed the lowest availability at 40.0%, while amoxicillin and salbutamol were also limited, at 43.3% and 46.7%, respectively (see Fig. 1). However, the inclusion of similar or therapeutically equivalent formulations for co-trimoxazole and diclofenac significantly improved availability for these medicines. Co-trimoxazole increased to 63.3%, and diclofenac reached the WHO-recommended target of 80%. As a result, the overall mean availability of essential medicines rose from 79.8% to 83.3%, highlighting the importance of considering alternative formulations in assessments of medicine accessAmong the fourteen WHO GCL medicines assessed, six were ≥ 80% available across all survey areas. In contrast, co-trimoxazole and salbutamol were not sufficiently available in any location. Notably, amoxicillin 500 mg was completely absent in Kavajë, while diclofenac and salbutamol were entirely unavailable in Tirana. The 80% national availability target was not reached in Durrës (75.7%), Kavajë (74.3%), and Tirana (72.9%), representing half of the surveyed locations. These figures underscore regional disparities in the availability of essential medicines (*see* Fig 2).

**Figure 1.**
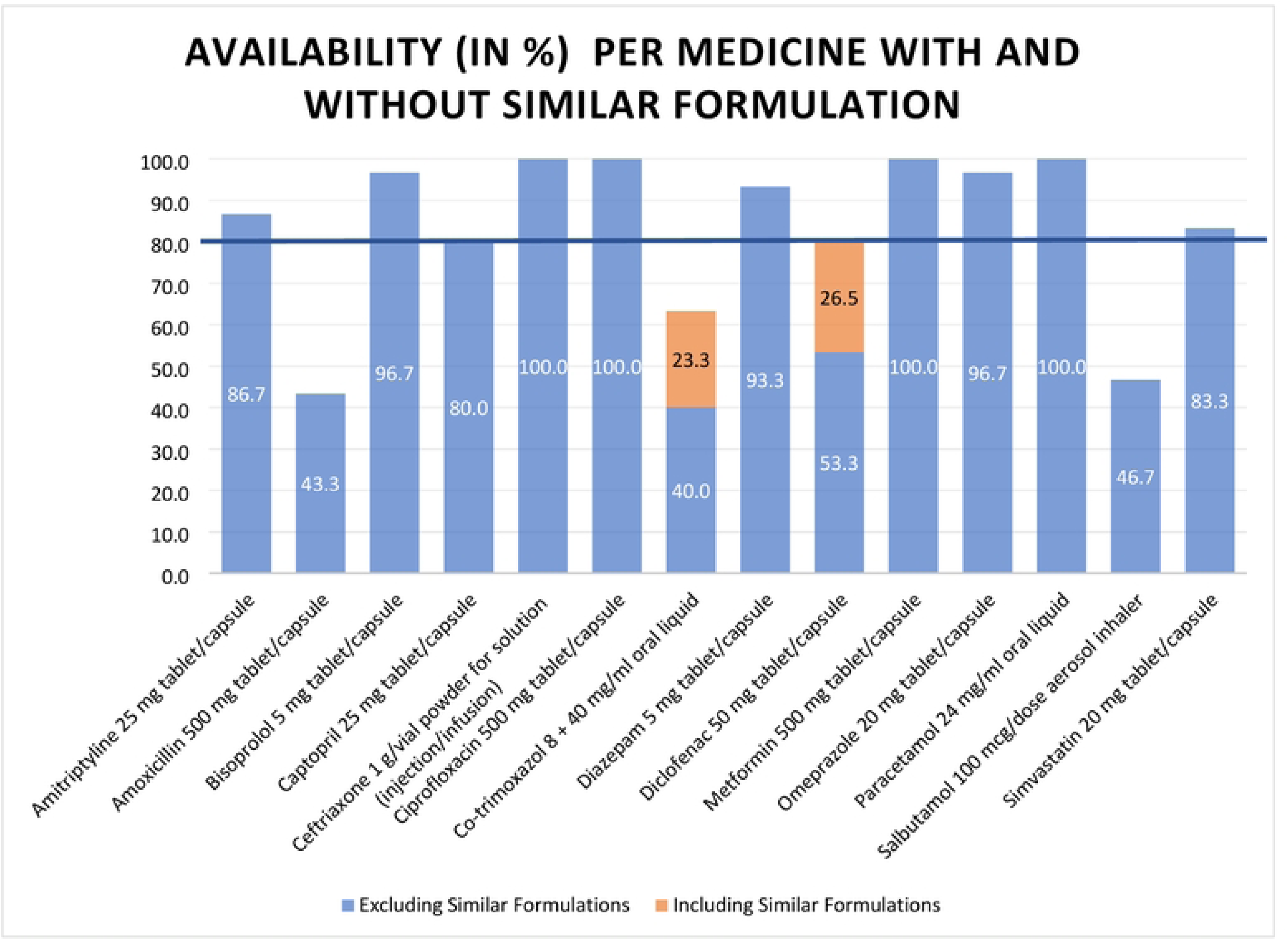
Availability (in %) per medicine included in the WHO Global Core List. ll1e blue line represents the 80% threshold of availability of essential medicines according 10 the WHO guidelines.

**Figure 2.**
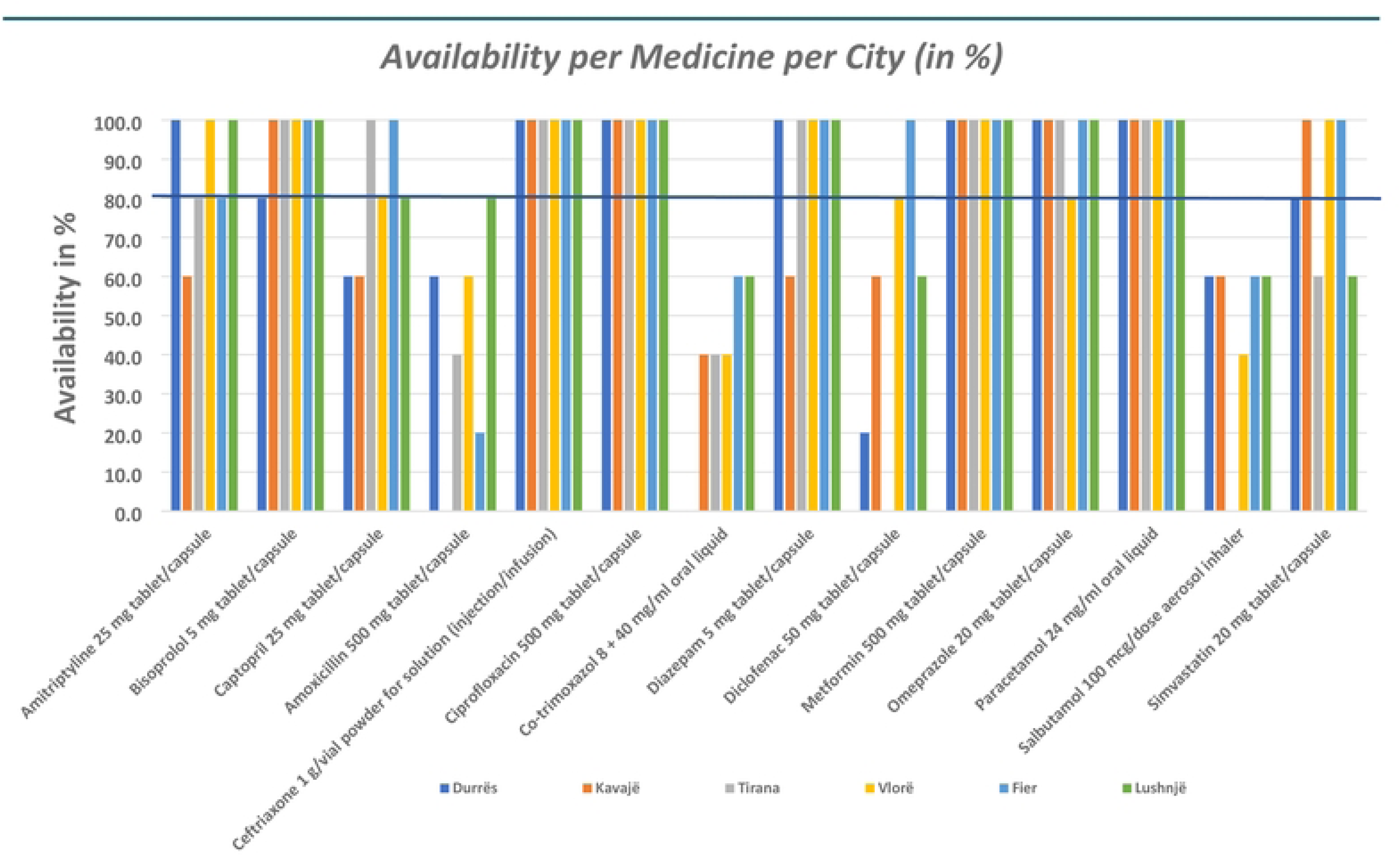
Availabilicy per medicine on the WHO Global Core List and percity. **The blue line** represents the **80% threshold** of availability of essential medicines. aocording to the WHO guidelines.

The availability of essential medicines showed clear disparities between OB and LPG formulations, as well as between contracted and uncontracted pharmacies (see Table 3). LPG formulations of core medicines were widely available, with ceftriaxone, ciprofloxacin, and metformin each reaching 100% availability. In comparison, OB formulations were found for only four medicines, and only metformin achieved an acceptable availability rate of 86.7%. Contracted pharmacies demonstrated higher mean availability (87.5%) compared to uncontracted ones (73.0%). Notably, medicines such as salbutamol (75.0% vs. 7.1%), diclofenac (75.0% vs. 28.6%), and simvastatin (100.0% vs. 64.3%) were significantly more available in contracted pharmacies, highlighting pronounced supply differences. In most cases where LPG formulations were lacking, OB alternatives were also not stocked. Diclofenac was the only exception, with OB versions more frequently available than LPG.

**Table 3.**
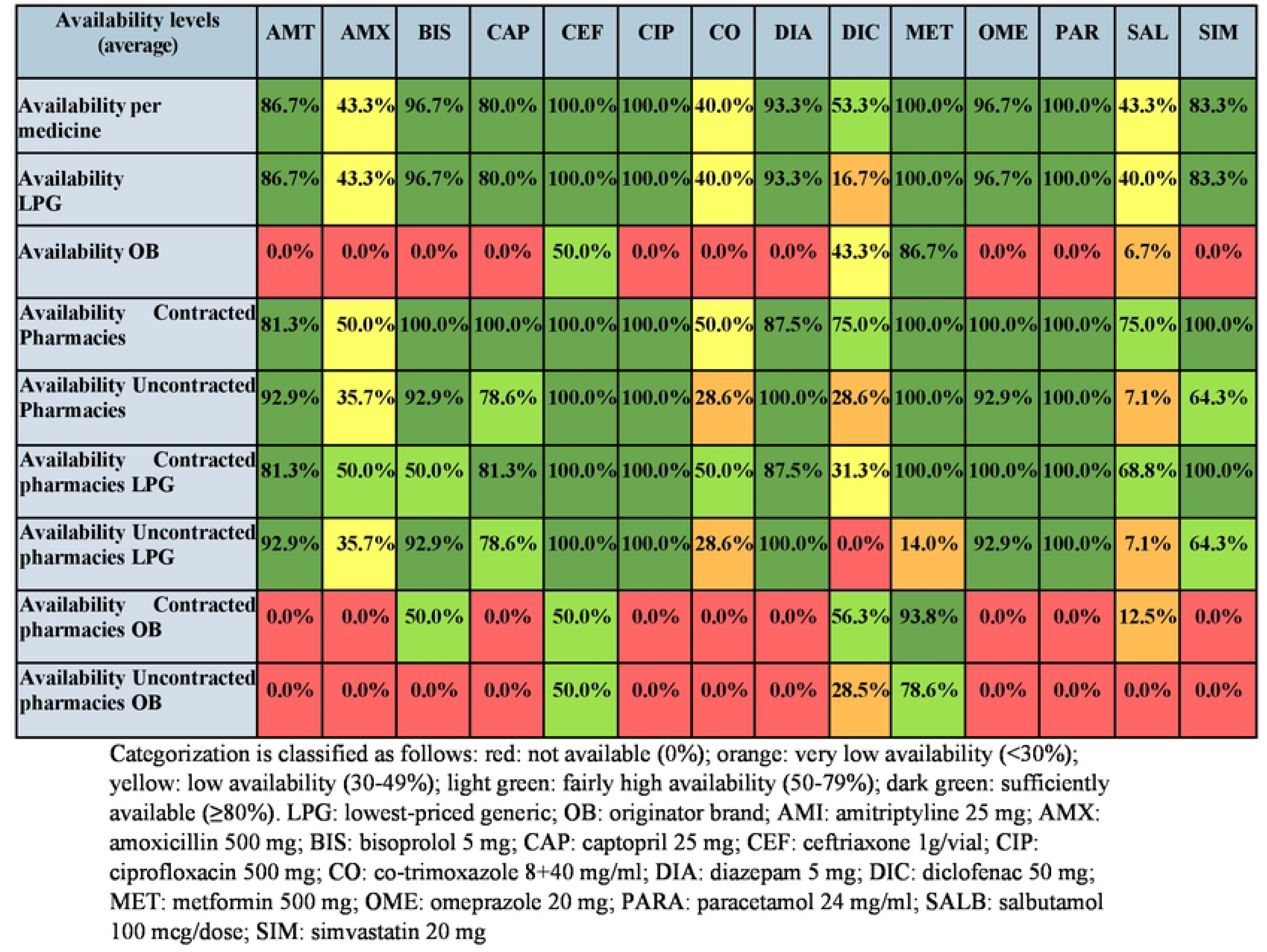
Availability of Surveyed Medicines by Brand Type (LPG/OB) and Phannacy Contract Status in Community Phannacies in Albania(%)

| Availability levels (average) | AMT | AMX | BIS | CAP | CEF | CIP | CO | DIA | DIC | MET | OME | PAR | SAL | SIM |
| --- | --- | --- | --- | --- | --- | --- | --- | --- | --- | --- | --- | --- | --- | --- |
| Availability per medicine | 86.7% | 43.3% | 96.7% | 80.0% | 100.0% | 100.0% | 40.0% | 93.3% | 53.3% | 100.0% | 96.7% | 100.0% | 43.3% | 83.3% |
| Availability LPG | 86.7% | 43.3% | 96.7% | 80.0% | 100.0% | 100.0% | 40.0% | 93.3% | 16.7% | 100.0% | 96.7% | 100.0% | 40.0% | 83.3% |
| Availability OB | 0.0% | 0.0% | 0.0% | 0.0% | 50.0% | 0.0% | 0.0% | 0.0% | 43.3% | 86.7% | 0.0% | 0.0% | 6.7% | 0.0% |
| Availability Contracted Pharmacies | 81.3% | 50.0% | 100.0% | 100.0% | 100.0% | 100.0% | 50.0% | 87.5% | 75.0% | 100.0% | 100.0% | 100.0% | 75.0% | 100.0% |
| Availability Uncontracted Pharmacies | 92.9% | 35.7% | 92.9% | 78.6% | 100.0% | 100.0% | 28.6% | 100.0% | 28.6% | 100.0% | 92.9% | 100.0% | 7.1% | 64.3% |
| Availability Contracted pharmacies LPG | 81.3% | 50.0% | 50.0% | 81.3% | 100.0% | 100.0% | 50.0% | 87.5% | 31.3% | 100.0% | 100.0% | 100.0% | 68.8% | 100.0% |
| Availability Uncontracted pharmacies LPG | 92.9% | 35.7% | 92.9% | 78.6% | 100.0% | 100.0% | 28.6% | 100.0% | 0.0% | 14.0% | 92.9% | 100.0% | 7.1% | 64.3% |
| Availability Contracted pharmacies OB | 0.0% | 0.0% | 50.0% | 0.0% | 50.0% | 0.0% | 0.0% | 0.0% | 56.3% | 93.8% | 0.0% | 0.0% | 12.5% | 0.0% |
| Availability Uncontracted pharmacies OB | 0.0% | 0.0% | 0.0% | 0.0% | 50.0% | 0.0% | 0.0% | 0.0% | 28.5% | 78.6% | 0.0% | 0.0% | 0.0% | 0.0% |

Categorization is classified as follows: red: not available (0%); orange: very low availability (<30%); yellow: low availability (30-49%); light green: fairly high availability (50-79%); dark green: sufficiently available ( $\geq 80\%$ ). LPG: lowest-priced generic; OB: originator brand; AMI: amitriptyline 25 mg; AMX: amoxicillin 500 mg; BIS: bisoprolol 5 mg; CAP: captopril 25 mg; CEF: ceftriaxone 1g/vial; CIP: ciprofloxacin 500 mg; CO: co-trimoxazole 8+40 mg/ml; DIA: diazepam 5 mg; DIC: diclofenac 50 mg; MET: metformin 500 mg; OME: omeprazole 20 mg; PARA: paracetamol 24 mg/ml; SALB: salbutamol 100 mcg/dose; SIM: simvastatin 20 mg

Stock-outs were reported in 25 of the 30 surveyed health facilities (83%), with a total of 58 individual stock-out incidents.

These varied in duration, with 31 medicines experiencing stock-outs exceeding one month, 21 lasting one month, and 4 lasting one week; in two cases, the duration was unspecified. The most frequently affected medicines were salbutamol (19 stock-outs), co-trimoxazole (n = 13), amoxicillin (n = 12), and diclofenac (n = 8), which were also among those most commonly reported as unavailable.

### Affordability

Following the WHO/HAI methodology, median unit prices were calculated for all surveyed medicines, along with MPRs relative to selected Balkan countries, EU member states, and international reference prices (see Table S3). Median prices varied considerably—for example, 1.14 ALL per dose of salbutamol and 813.50 ALL per vial of ceftriaxone—possibly reflecting differences in dosage form, therapeutic category, and procurement context. Several medicines, including diclofenac (MPR = 11.35) and ceftriaxone (MPR = 15.08), showed particularly high MPRs compared to international reference prices, raising concerns about affordability and pricing efficiency.

#### Affordability as per Lowest Paid Government Worker approach

According to the LPWG approach, all core medicines were considered affordable except for ceftriaxone 1 g/vial (see Table 4). Diazepam and simvastatin consistently had very low day’s wage values of 0.01 and 0.10, respectively. Co-trimoxazole had daily’s wage values of 0.08 in contracted pharmacies, compared to 0.14 in uncontracted pharmacies.

**Table 4.**
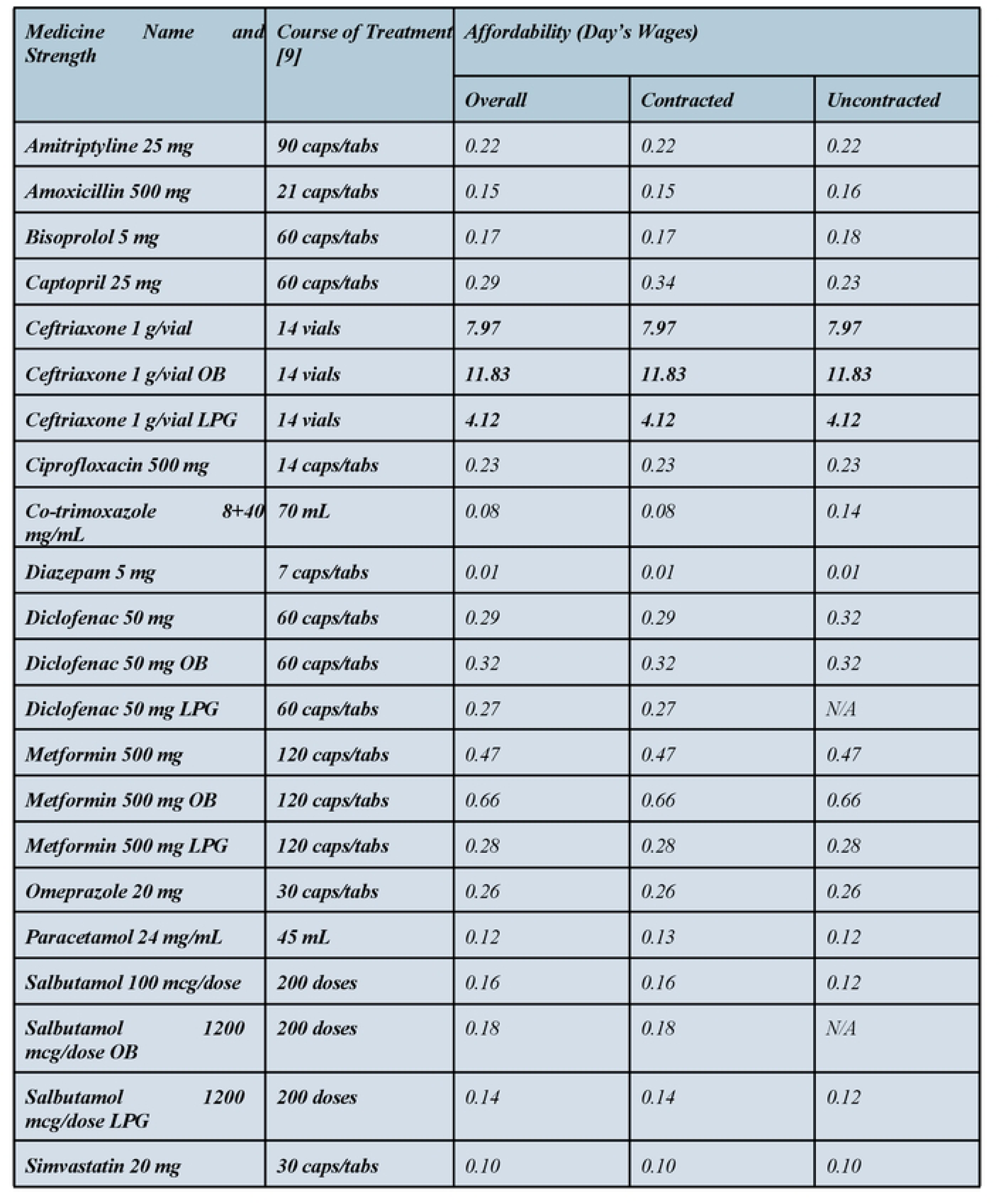
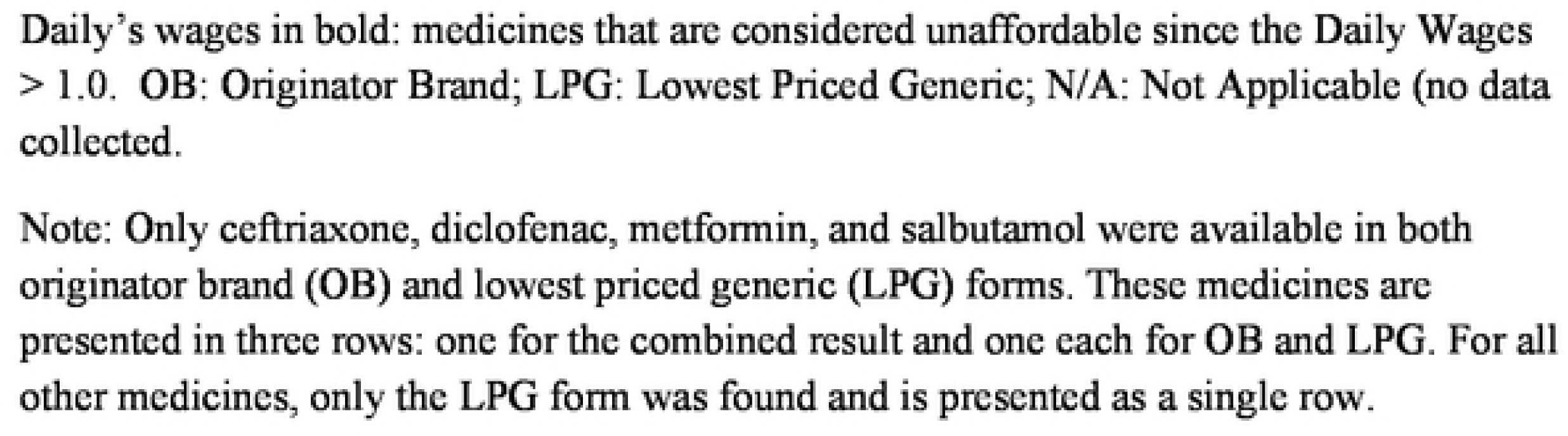
Overall affordability, affordability in contracted pharmacies and affordability in uncontracted pharmacies (median prices), using the LPWG approach.

| *Medicine Name and Strength* | *Course of Treatment [9]* | *Affordability (Day's Wages)* |  |  |
| --- | --- | --- | --- | --- |
|  |  | *Overall* | *Contracted* | *Uncontracted* |
| *Amitriptyline 25 mg* | *90 caps/tabs* | *0.22* | *0.22* | *0.22* |
| *Amoxicillin 500 mg* | *21 caps/tabs* | *0.15* | *0.15* | *0.16* |
| *Bisoprolol 5 mg* | *60 caps/tabs* | *0.17* | *0.17* | *0.18* |
| *Captopril 25 mg* | *60 caps/tabs* | *0.29* | *0.34* | *0.23* |
| *Ceftriaxone 1 g/vial* | *14 vials* | *7.97* | *7.97* | *7.97* |
| *Ceftriaxone 1 g/vial OB* | *14 vials* | *11.83* | *11.83* | *11.83* |
| *Ceftriaxone 1 g/vial LPG* | *14 vials* | *4.12* | *4.12* | *4.12* |
| *Ciprofloxacin 500 mg* | *14 caps/tabs* | *0.23* | *0.23* | *0.23* |
| *Co-trimoxazole 8+40 mg/mL* | *70 mL* | *0.08* | *0.08* | *0.14* |
| *Diazepam 5 mg* | *7 caps/tabs* | *0.01* | *0.01* | *0.01* |
| *Diclofenac 50 mg* | *60 caps/tabs* | *0.29* | *0.29* | *0.32* |
| *Diclofenac 50 mg OB* | *60 caps/tabs* | *0.32* | *0.32* | *0.32* |
| *Diclofenac 50 mg LPG* | *60 caps/tabs* | *0.27* | *0.27* | *N/A* |
| *Metformin 500 mg* | *120 caps/tabs* | *0.47* | *0.47* | *0.47* |
| *Metformin 500 mg OB* | *120 caps/tabs* | *0.66* | *0.66* | *0.66* |
| *Metformin 500 mg LPG* | *120 caps/tabs* | *0.28* | *0.28* | *0.28* |
| *Omeprazole 20 mg* | *30 caps/tabs* | *0.26* | *0.26* | *0.26* |
| *Paracetamol 24 mg/mL* | *45 mL* | *0.12* | *0.13* | *0.12* |
| *Salbutamol 100 mcg/dose* | *200 doses* | *0.16* | *0.16* | *0.12* |
| *Salbutamol 1200 mcg/dose OB* | *200 doses* | *0.18* | *0.18* | *N/A* |
| *Salbutamol 1200 mcg/dose LPG* | *200 doses* | *0.14* | *0.14* | *0.12* |
| *Simvastatin 20 mg* | *30 caps/tabs* | *0.10* | *0.10* | *0.10* |

Daily's wages in bold: medicines that are considered unaffordable since the Daily Wages > 1.0. OB: Originator Brand; LPG: Lowest Priced Generic; N/A: Not Applicable (no data collected).
Note: Only ceftriaxone, diclofenac, metformin, and salbutamol were available in both originator brand (OB) and lowest priced generic (LPG) forms. These medicines are presented in three rows: one for the combined result and one each for OB and LPG. For all other medicines, only the LPG form was found and is presented as a single row.

There were no differences found between affordability in contracted and uncontracted pharmacies concerning those treatments exceeding the 1-day threshold. Diclofenac 50 mg LPG and salbutamol 100 mcg/dose were not available in contracted pharmacies and were therefore excluded from the analysis.

#### Affordability using the Median Price Ratio

The comparative analysis of medicine prices revealed that, for nearly all core medicines, unit prices in Albania and the corresponding IRPs were lower than the average prices observed in both the selected Balkan and EU reference countries (*see* Fig 3). This trend was particularly pronounced for commonly used medicines such as amoxicillin, ciprofloxacin, diazepam, diclofenac, omeprazole, and simvastatin.

**Figure 3.**
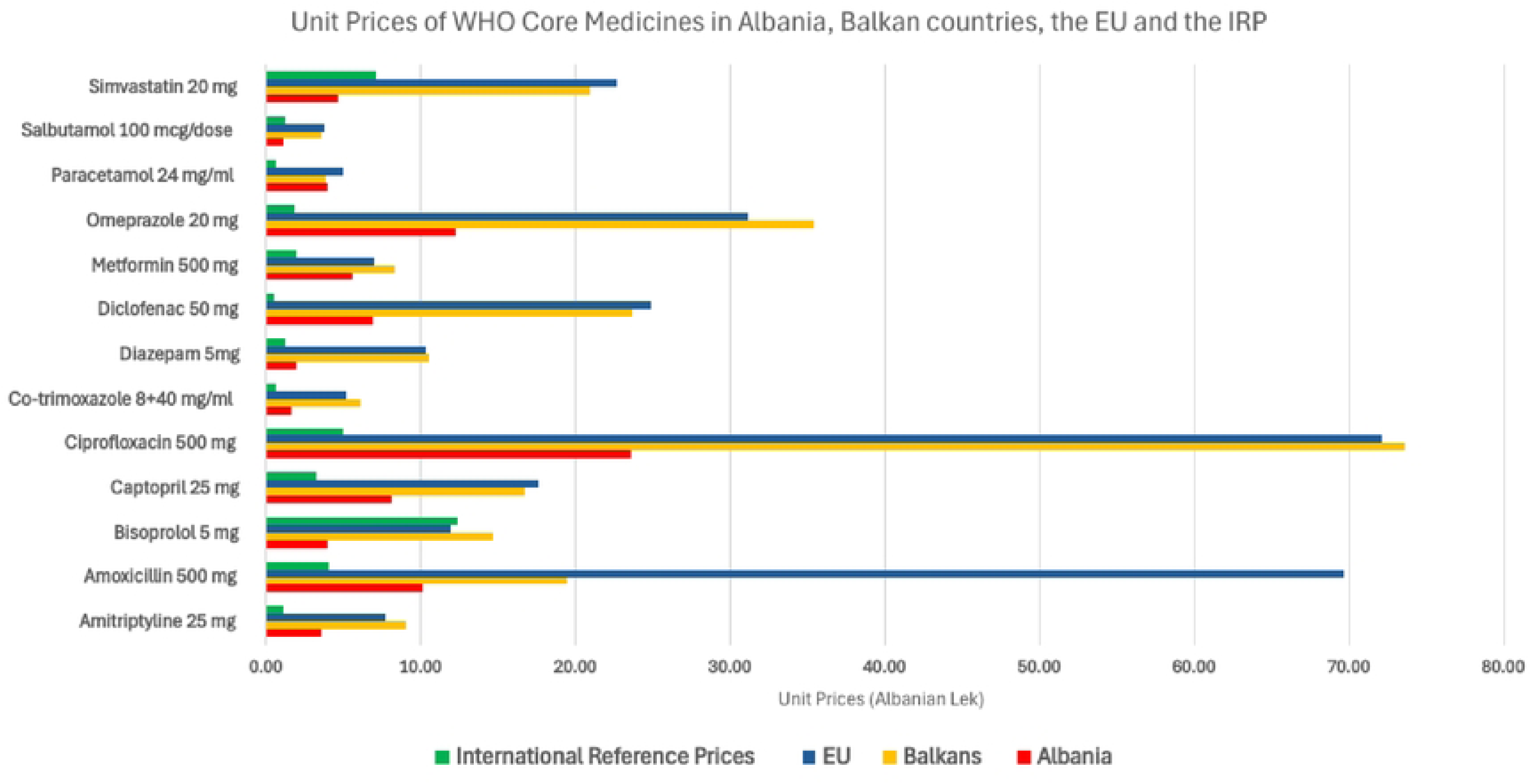
Unit prices of WHO Core Medicines (excluding ceftriaxone) in Albania, reference regions (Balkans, EU) compared to the international reference prices.

Further insight is provided by the MPR values presented in Table 5. MPRs confirm that amoxicillin and diazepam are up to five times more affordable in Albania than in EU countries, with a similar level of affordability observed for diazepam in Balkan countries. When benchmarked against International Reference Prices (IRPs), only three core medicines (bisoprolol, salbutamol and simvastatin) were found to be more affordable in Albania.. In contrast, five medicines recorded MPRs exceeding 4.0, indicating prices significantly higher than international reference prices. These findings paint a mixed pricing landscape in Albania, where certain essential medicines are competitively priced, while others remain relatively expensive. To improve clarity, ceftriaxone was excluded from the visual comparison due to its outlier status; however, its unit price values are presented separately: IRP 42.26, EU countries 588.38, Balkan countries 897.18, and Albania 813.50 (*see* Table S3).

**Table 5.**
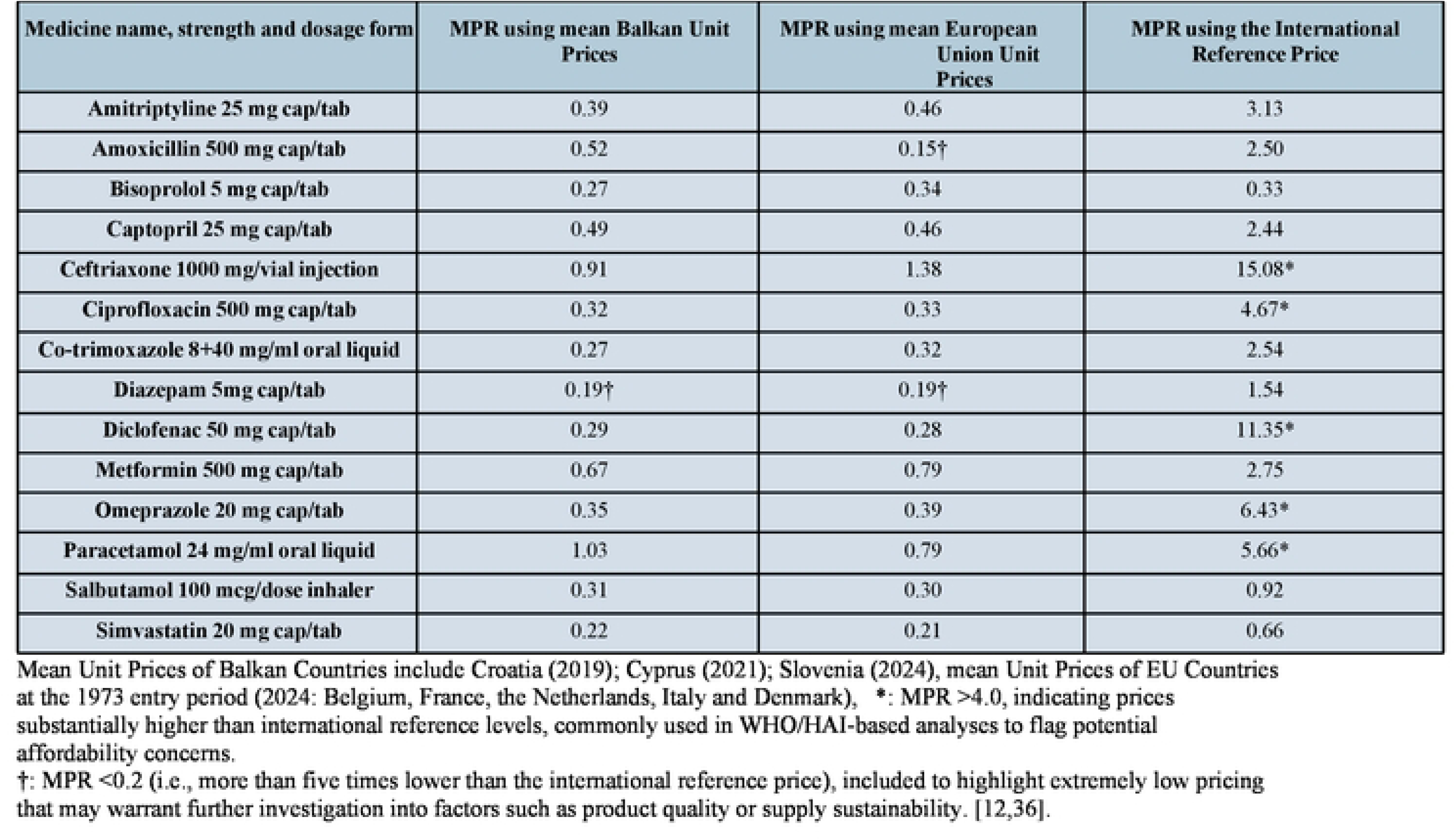
Median Price Ratios using Balkan and EU Reference countries and the International Reference Price.

#### Accessibility

An assessment of overall accessibility of core medicines in Albania was undertaken by combining data on availability and affordability, the latter calculated using the LPGW method (see Fig 4). The analysis revealed that limited accessibility was primarily driven by low availability, which affected 4 out of the 14 core medicines surveyed. In contrast, affordability was not a widespread issue and was identified only for ceftriaxone 1 g/vial treatments, where the cost exceeded the accepted affordability threshold.

**Figure 4.**
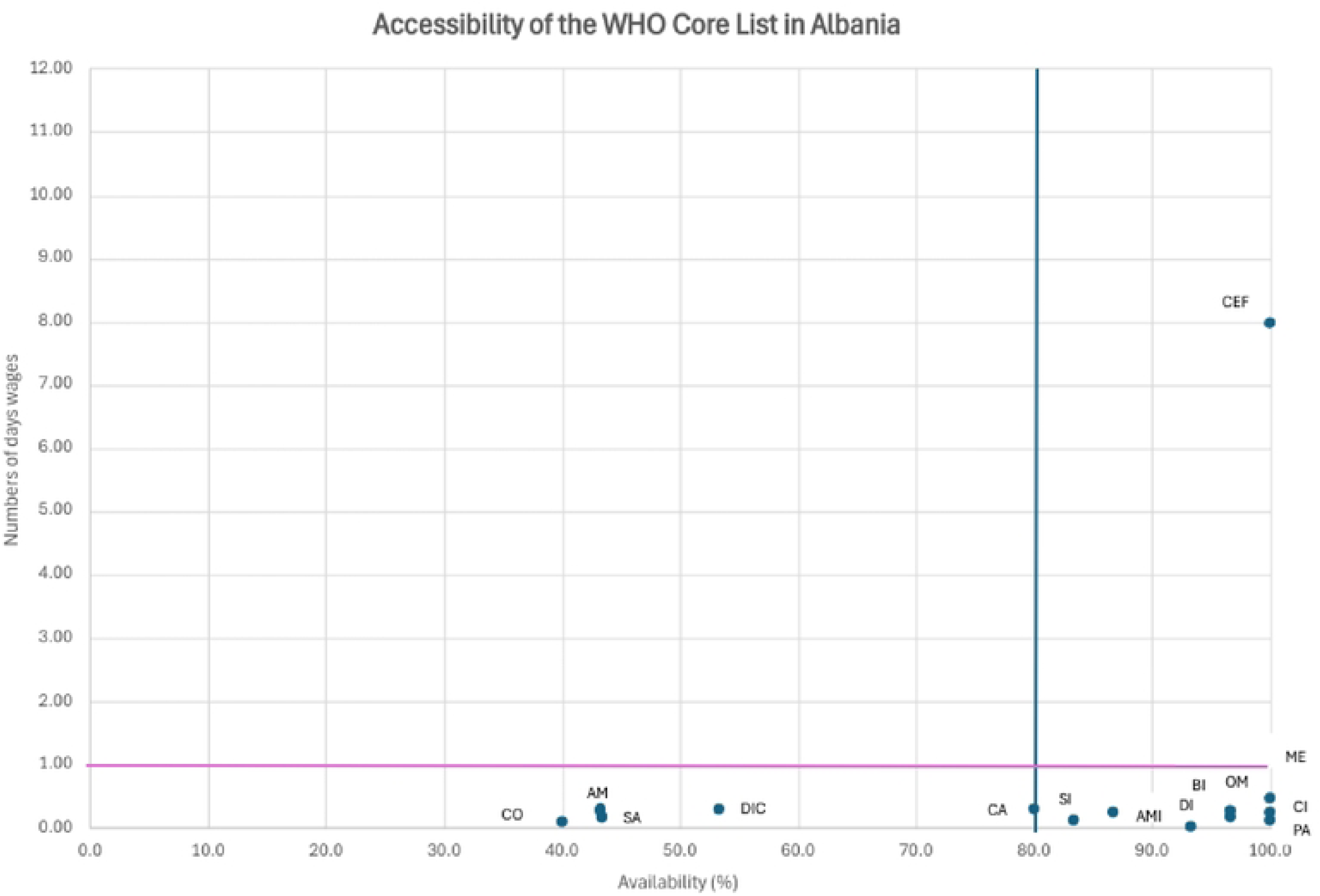
Overall accessibility of the WHO Global Core List using the overall availability (in %) and the median LPGW approach. The blue line represents the 80% threshold of availability of EM’s according to the WHO guidelines. The pink line represents the I-day threshold related to the Daily Wages needed towork to afford the medicine for the LPGW. AMI: amitriptyline 25 mg; AM: amoxicillin 500 mg; BI: bisoprolol 5 mg; CA: captopril 25 mg; CEF: ceftriaxone lg/vial; CJ: ciprofloxacin 500 mg; CO: co-trimoxazol 8+40 mg/ml; DI: diazepam 5 mg; DIC: diclofenac 50 mg; ME: metformin 500 mg; OM: omcprazole 20 mg; PA: paracetamol 24 mg/ml; SA: salbutamol 100 mcg/dose; SI: simvastatin 20 mg.

Based on the combined criteria, 9 out of 14 core medicines were classified as accessible, resulting in an overall accessibility level of 64%. This falls short of the widely accepted benchmark of 80% accessibility for essential medicines.

## Discussion

Access to essential medicines, particularly those listed in the WHO Global Core List (GCL), is central to the right to health and critical for reducing the burden of non-communicable diseases (NCDs). In 2013, the WHO set a target of 80% availability of affordable essential NCD medicines and technologies by 2025 to support a 25% reduction in premature NCD mortality [37]. This goal was reinforced by Sustainable Development Goal (SDG) target 3.4, which aims to reduce premature NCD mortality by one third by 2030 through prevention and treatment [38]. To support progress, the WHO launched the *Implementation Roadmap 2023–2030*, which outlines strategies to expand access to essential medicines and strengthen health systems [37].

In this context, the present Albanian study assessed the availability, affordability, and pricing of core essential NCD medicines. Of the 14 core medicines surveyed, four were unavailable at the time of assessment. Affordability analysis based on the LPGW method showed that all medicines except ceftriaxone were considered affordable. However, pricing analysis using the Median Price Ratio (MPR) method revealed that only 3 out of 14 medicines were priced below the international reference price (IRP), while five had MPRs exceeding the affordability benchmark of 4.0. With an overall accessibility rate of 64%, the study indicates that Albania did not meet the WHO’s 2025 target.

The average availability of medicines from the GCL in Albania was 79.8%, with regional disparities observed. LPGs were 2.5 times more available than OBs. The availability of medicines from the GCL in the WHO Europe region has been assessed in Kazakhstan, Moldova and Russia (upper-middle income countries) and Uzbekistan, Tajikistan and Ukraine (lower-middle income countries) *(see* Table S5). Moldova, comparable to Albania in population size and healthcare challenges, found a mean overall availability of 67.9%, compared to 70.1% and 70.9% in the three upper-middle income countries and lower-middle income countries, respectively [12,40].

Notably, several antibiotics from the Access group, which the World Health Organization (WHO) defines as first-or second-line treatments that should be widely available, affordable, and have a lower potential for resistance, were among the medicines found to be unavailable. This included commonly used antibiotics such as amoxicillin and co-trimoxazole. This lack of availability increases reliance on Watch group antibiotics, which are recommended only for specific, limited indications due to their higher potential to cause antimicrobial resistance.

Unavailability of access group antibiotics may thus contribute to antimicrobial resistance (AMR) [41]. One potential solution is the development and implementation of a comprehensive National Action Plan (NAP) on AMR. While Albania has a draft version of this NAP, delays in its approval contribute to the continued use of Watch antibiotics. The NAP aims to promote rational prescribing, strengthen surveillance, and support the use and availability of Access antibiotics through better stewardship and procurement alignment [9].

Salbutamol, an essential medicine for asthma treatment, was also unavailable in several uncontracted pharmacies in Albania, reflecting a broader issue observed across Europe [42–45]. Shortages have been reported in multiple EU countries, largely due to increased demand and global supply disruptions [44]. A key vulnerability lies in the concentration of active pharmaceutical ingredient (API) production in a small number of Asian manufacturers [45]. To address these challenges, the European Commission has proposed the Critical Medicines Act, which aims to diversify API sources and strengthen EU pharmaceutical resilience [48].

Only ceftriaxone, both its OB and LPG, was unaffordable by LPGW standards. On average, LPG medicines were 1.9 times more affordable and 2.5 times more available than OB medicines.

Retail medicine prices in Albania were relatively high, with an average MPR of 4.3 based on the WHO/HAI methodology using 2015 MSH IRPs adjusted for inflation. Only 3 of 14 core medicines had MPRs below 1.0, while five exceeded 4.0, suggesting pricing inefficiencies [12,16,34,40]. It should be noted that IRPs reflect global procurement prices and are not corrected for national purchasing power parity (PPP), which is methodologically complex and rarely done [47].

Generic medicines were generally more affordable and available than originator brands. While most core medicines were priced lower in Albania than in selected EU and Balkan countries, cross-country comparisons were complicated by differences in regulatory frameworks, price levels (wholesale vs. retail), and reimbursement structures [32,52]. Notably, the only medicine deemed unaffordable (ceftriaxone) using the LPGW approach was not included in the Albanian Medicines Reimbursement List (AMHIF), highlighting a gap in financial protection. These findings underscore the need for further investigation into procurement pricing transparency, supply chain mark-ups, and the role of reimbursement mechanisms in ensuring medicine affordability and access in Albania.

### Strengths and Limitations

This study has several strengths. It used an adapted version of the WHO/HAI methodology, a well-established and reliable tool for measuring the availability and affordability of medicines. This allows the results to be compared with studies from other countries. By focusing on medicines from the GCL, the study supports international efforts to monitor access to essential medicines. An extra analysis on how long medicines were out of stock and similar formulations was included, helping to better understand medicine availability in Albania. The results are also important for health policy, especially since Albania does not have its own Essential Medicines List. Finally, the study is one of the few of its kind in South-East Europe, and it could help guide future research and the creation of a regional medicine list that better reflects the health needs of this area.

However, there are also some limitations, with several stemming from inherent constraints of the WHO/HAI methodology. First, medicine availability was measured only on the day of the survey. This means the study did not capture how often or how long medicines were out of stock. Although an additional analysis was conducted, data may still be incomplete due to inconsistent reporting or pharmacists’ reluctance to repeat information. Only one strength and form per medicine was assessed; however, similar therapeutic alternatives were included to provide a broader view of availability.

Third, affordability was assessed using a general income estimate based on the lowest-paid government worker (LPGW), which does not consider other living costs or patients using multiple medicines. As a result, treatments may appear more affordable than they actually are in practice. Additionally, the analysis relied on outdated international reference prices (IRPs) from the 2015 MSH price guide, which may limit the accuracy of present-day price comparisons. As highlighted by Wirtz et al. [49], transparent and current price data are essential for equitable access and sound pharmaceutical policy. Outdated benchmarks may therefore compromise the validity of cross-country comparisons.

Furthermore, the study focused only on the 14 medicines included in the WHO Global Core List, without considering those from regional or supplementary lists that may better reflect national disease burdens. Lastly, the study did not assess medicine acceptability, including patient preferences, adherence, and perceptions of quality, which are critical dimensions of access and may significantly influence medicine use and treatment outcomes. Future research should address these factors to provide a more comprehensive understanding of access to essential medicines in Albania.

## Conclusion

This study provides insights into the accessibility of the WHO Global Core List of essential medicines in Albania. Availability issues, particularly with antibiotics and asthma medications, highlight the need for better policy on rational use and other interventions along the pharmaceutical value chain. While medicines were generally more affordable in Albania compared to Europe, affordability concerns arose with ceftriaxone. These insights should inform Albania’s health policy and align with EU health protection goals, while underscoring the importance of regular accessibility studies to support the achievement of SDG 3.b.3 by 2030.

## Data Availability

All relevant data are within the manuscript and its Supporting Information files.

N\A

## Abbreviations

ALL: Albanian Lek (currency)
AMHIF: Albanian Mandatory Health Insurance Fund
CPI: Consumer Price Index
EU: European Union
GCL: Global Core List
GDP: Gross Domestic Product
HAI: Health Action International
IRP: International Reference Price
LMIC: Low– and Middle-Income Countries
LPG: Lowest Priced Generic
LPGW: Lowest-Paid Government Worker
MPR: Median Price Ratio
OB: Originator Brand
QSUT: University Hospital Center “Mother Teresa” (Tirana)
SDG: Sustainable Development Goal
UMIC: Upper-Middle-Income Country
WHO: World Health Organization

## References

1. World Health Organization. Equitable access to essential medicines: a framework for collective action. Geneva: World Health Organization; 2004.

2. Penchansky R, Miralles M, Walkowiak H, Boesen D, Burn R, Chalker JM. Defining and measuring access to essential drugs, vaccines, and health commodities: report of the WHO-MSH consultative meeting. Arlington (VA): Centre for Pharmaceutical Management; 2003.

3. World Health Organization. Essential medicines [Internet]. Geneva: World Health Organization; 2023 [cited 2025 May 26]. Available from: https://www.who.int/news-room/fact-sheets/detail/essential-medicines

4. Institute of Statistics (INSTAT). Population | Demography and social indicators. Population of Albania on 1st January 2025 [Internet]. Tirana: INSTAT; [cited 2025 Jul 10]. Available from: https://www.instat.gov.al/en/themes/demography-and-social-indicators/population/#tab2

5. The Global Economy. Albania: Health spending per capita. TheGlobalEconomy.com; 2021 [cited 2025 Jul 10]. Available from: https://www.theglobaleconomy.com/Albania/Health_spending_per_capita

6. Petro E, Perumal-Pillay V, Mantel-Teeuwisse AK, van den Ham HA, Suleman F. Evaluation of alignment of the reimbursement medicines list for children in Albania with the WHO essential medicines list for children. J Pharm Policy Pract. 2023;17(1):2290100.

7. European Commission. Chapters of Acquis [Internet]. [cited 2024 Apr 26]. Available from: https://neighbourhood-enlargement.ec.europa.eu/enlargement-policy/conditions-membership/chaptersacquis_en

8. Institute of Public Health. Non-communicable diseases (NCD) in Albania 2020. Tirana: Institute of Public Health; 2020. Available from: https://www.ishp.gov.al/wp-content/uploads/2021/06/NCD-Albania-summary-2020.pdfishp.gov.al

9. Hoxha I, Malaj A, Kraja B, et al. 11-Year Trend in Antibiotic Consumption in a South-Eastern European Country: The Situation in Albania and the Implications for the Future. Antibiotics (Basel). 2023;12(1):89. doi:10.3390/antibiotics12010089ResearchGate+1PubMedCentral+1

10. Beci A, Belishova A, Kola E. *Kush* paguan: Financimi i shërbimeve shëndetësore në Shqipëri [Internet]. Tirana (Albania): 2015 [cited 2024 Apr 9]. Available from: https://fsdksh.gov.al/wp-content/uploads/2019/11/Kush_Paguan_web.pdf

11. Fondi i Sigurimit të Detyrueshëm të Kujdesit Shëndetësor. Lista e barnave të rimbursueshme [Internet]. [cited 2025 Apr 29]. Available from: https://fsdksh.gov.al/project/lista-e-barnave/

12. World Health Organization, Health Action International. Measuring medicine prices, availability, affordability and price components. 2nd ed. Geneva: World Health Organization; 2008. Available from: https://iris.who.int/handle/10665/70013

13. World Bank. Population, total – Albania [Internet]. Washington, DC: World Bank; [cited 2024 Apr 28]. Available from: https://data.worldbank.org/indicator/SP.POP.TOTL?end=2020&locations=AL&start=1990&view=chart

14. World Health Organization. The Global Health Observatory: Explore a world of health data [Internet]. Geneva: WHO; 2023 [cited 2024 May 1]. Available from: https://www.who.int/data/gho/data/indicators/indicators-index

15. World Bank. GDP per capita (current US$) [Internet]. Washington, DC: World Bank; [cited 2024 Apr 29]. Available from: https://data.worldbank.org/indicator/NY.GDP.PCAP.CD

16. European Commission. Healthcare expenditure statistics by function, provider and financing scheme [Internet]. Eurostat Statistics Explained. 2024 [cited 2025 Apr 29]. Available from: https://ec.europa.eu/eurostat/statistics-explained/index.php?title=Healthcare_expenditure_statistics_by_function,_provider_and_financing_scheme

17. World Health Organization. Life expectancy at birth [Internet]. Geneva: World Health Organization; [cited 2024 Apr 29]. Available from: https://www.who.int/data/gho/data/indicators/indicator-details/GHO/life-expectancy-at-birth-%28years%29

18. World Bank. Poverty headcount ratio at $3.65 a day (2011 PPP) (% of population) [Internet]. Washington, DC: World Bank; [cited 2024 Apr 29]. Available from: https://data.worldbank.org/indicator/SI.POV.DDAY

19. European Commission. At risk of poverty or social exclusion (AROPE) [Internet]. Eurostat Statistics Explained. 2024 [cited 2025 Apr 29]. Available from: https://ec.europa.eu/eurostat/statistics-explained/index.php?title=Glossary:At_risk_of_poverty_or_social_exclusion_(AROPE)

20. Trading Economics. Albania Unemployment Rate. [Internet]. 2024 [cited 2025 Apr 29]. Available from: https://tradingeconomics.com/albania/unemployment-rate

21. YCharts. European Union Unemployment Rate (I:EUUR). [Internet]. 2025 [cited 2025 Apr 29]. Available from: https://ycharts.com/indicators/european_unemployment_rate_YCharts+3

22. World Bank. Mortality rate, neonatal (per 1,000 live births) [Internet]. Washington, D.C.: World Bank; [cited 2025 Apr 29]. Available from: https://data.worldbank.org/indicator/SH.DYN.NMRT

23. World Health Organization. Crude death rate (deaths per 1,000 population) [Internet]. Geneva: World Health Organization; [cited 2025 Apr 29]. Available from: https://data.who.int/indicator/GHO/crude-death-rate-deaths-per-1000-population

24. World Health Organization. Density of health workers [Internet]. Geneva: World Health Organization; [cited 2024 Apr 29]. Available from: https://www.who.int/data/gho/data/themes/health-workforce

25. World Health Organization. UHC service coverage index [Internet]. Geneva: WHO; 2024 [cited 2025 Apr 29]. Available from: https://data.who.int/indicators/i/3805B1E/9A706FD

26. World Health Organization. Diabetes prevalence [Internet]. Geneva: World Health Organization; [cited 2024 Apr 29]. Available from: https://www.who.int/data/gho/data/themes/topics/topic-details/GHO/diabetes

27. Global Burden of Disease Study. Probability of dying from NCDs between ages 30 and 70 [Internet]. Seattle, WA: Institute for Health Metrics and Evaluation (IHME); [cited 2024 Apr 29]. Available from: http://ghdx.healthdata.org/gbd-results-tool

28. KONTRATAT ME DHËNËSIT E SHËRBIMIT PARËSOR. [Internet]. Available from: https://fsdksh.gov.al/qendra-shendetesore/. Last Accessed on: 13-05-2024.

29. Agjencia Kombetare e Barnave dhe Pajisjeve Mjekesore. Regjistri i Barnave. [Internet]. Available from: https://akbpm.gov.al/. Last Accessed on: 05-05-2024.

30. KoboToolbox [Internet]. Available from: https://www.kobotoolbox.org. Accessed 2024 Aug 5.

31. Institute of Statistics. Wage Statistics, Q4 – 2024 [Internet]. Tirana: Institute of Statistics; 2025 Mar 11 [cited 2025 Apr 9]. Available from: https://www.instat.gov.al/media/14985/wage-statistics-q4-2024.pdf

32. World Health Organization. Medicine price information sources [Internet]. Geneva: WHO; [cited 2024 Apr 24]. Available from: https://www.who.int/teams/health-product-and-policy-standards/medicines-selection-ip-and-affordability/affordability-pricing/med-price-info-source

33. International Monetary Fund. Cross-country indexes, year-over-year change [Internet]. Washington (DC): IMF; [cited 2024 May 20]. Available from: https://data.imf.org/regular.aspx?key=61015894

34. World Health Organization. Median consumer price ratio of selected medicines [Internet]. Geneva: WHO; [cited 2024 Jun 23]. Available from: https://www.who.int/data/gho/indicator-metadata-registry/imr-details/11

35. YCharts. US Consumer Price Index (I:USCPI) [Internet]. [cited 2024 May 20]. Available from: https://ycharts.com/indicators/us_consumer_price_index

36. World Health Organization. Source Balkan and EU countries [Internet]. Geneva: WHO; [cited 2025 Apr 10]. Available from: https://www.who.int/teams/health-product-and-policy-standards/medicines-selection-ip-and-affordability/affordability-pricing/med-price-info-source

37. World Health Organization. Global Action Plan for the Prevention and Control of Noncommunicable Diseases 2013–2020. Geneva: WHO; 2013.

38. United Nations. Transforming our world: the 2030 Agenda for Sustainable Development. New York: UN; 2015.

39. World Health Organization. NCD Implementation Roadmap 2023–2030: to support the achievement of target 3.4 of the Sustainable Development Goals. Geneva: WHO; 2023.

40. Deng J, Mayai AT, Kayitare E, Atwine R, Akunne OO, et al. Assessment of prices, availability and affordability of essential medicines in Juba County, South Sudan. J Pharm Policy Pract. 2023;16:172. 10.1186/s40545-023-00675-5

41. BD 2019 Antimicrobial Resistance Collaborators. Global mortality associated with 33 bacterial pathogens in 2019: a systematic analysis for the Global Burden of Disease Study 2019. Lancet. 2022 Dec;400(10369):2221–2248.

42. Mahase E. Doctors are told to ration salbutamol amid shortage. BMJ. 2024;384.

43. Drug Shortages Canada. Drug Shortage Report for VENTOLIN HFA [Internet]. [cited 2024 May 30]. Available from: https://www.drugshortagescanada.ca/shortage/208287

44. KNMP Farmanco. Salbutamol aerosol 100 µg/do [Internet]. [cited 2024 May 30]. Available from: https://farmanco.knmp.nl/geneesmiddel/3763.html

45. European Medicines Agency. Meeting summary – Medicine Shortages SPOC Working Party, 14 March 2024 [Internet]. Amsterdam: EMA; 2024 Mar. Available from: https://www.ema.europa.eu/en/documents/minutes/meeting-summary-medicine-shortages-spoc-working-party-14-march-2024_en.pdf

46. Euronews. Which critical medicines are in short supply in the European Union? Euronews. 2025 Mar 11. Available from: https://www.euronews.com/health/2025/03/11/which-critical-medicines-are-in-short-supply-in-the-european-union

47. European Fine Chemicals Group. Medicine shortages in Europe: causes and solutions. Executive Summary by IQVIA for EFCG. Brussels: EFCG; 2020. Available from: https://efcg.cefic.org/wp-content/uploads/2021/06/20201211_IQVIA-for-EFCG_Executive-summary.pdf

48. European Commission. Commission proposes Critical Medicines Act to bolster supply of critical medicines in the EU. Brussels: European Commission; 2025 Mar 11. Available from: https://ireland.representation.ec.europa.eu/news-and-events/news/commission-proposes-critical-medicines-act-bolster-supply-critical-medicines-eu-2025-03-11_en

49. Wirtz VJ, Hogerzeil HV, Gray AL, Bigdeli M, Joncheere CP, et al. Essential medicines for universal health coverage. The Lancet. February 2017;389(10067):403–476

50. European Health Management Association. Health Management in South-Eastern Europe: Challenges and Opportunities [internet]. Brussels: European Health Management Association; 2022 Dec. Available from: https://ehma.org/app/uploads/2022/12/Health-Management-in-South-Eastern-Europe-challenges-and-opportunities.pdf.

51. Yang H, Dib HH, Zhu M, Qi G, Zhang X. Prices, availability and affordability of essential medicines in rural areas of Hubei Province, China. Health Policy and Planning. 2010 May;25(3):219–229.

52. Fédération Européenne d’Associations et d’Industries Pharmaceutiques. Principles for application of international reference pricing systems [internet]. Brussels: Fédération Européenne d’Associations et d’Industries Pharmaceutiques; 2014. Available from: https://www.efpia.eu/media/15406/efpia-position-paper-principles-for-application-of-international-reference-pricing-systems-june-2014.pdf.

